# Dissecting heart age using cardiac magnetic resonance videos, electrocardiograms, biobanks, and deep learning

**DOI:** 10.1101/2021.06.09.21258645

**Authors:** Alan Le Goallec, Jean-Baptiste Prost, Sasha Collin, Samuel Diai, Théo Vincent, Chirag J. Patel

## Abstract

Heart disease is the first cause of death after age 65 and, with the world population aging, its prevalence is expected to starkly increase. We used deep learning to build a heart age predictor on 45,000 heart magnetic resonance videos [MRI] and electrocardiograms [ECG] from the UK Biobank cohort (age range 45-81 years). We predicted age with a root mean squared error [RMSE] of 2.81±0.02 years (R-Squared=85.6±0.2%) and found that accelerated heart aging is heritable at more than 35%. MRI-based anatomical features predicted age better than ECG-based electro-physiological features (RMSE=2.89±0.02 years vs. 6.09±.0.02 years), and heart anatomical and electrical aging are weakly correlated (Pearson correlation=.249±.002). Our attention maps highlighted the aorta, the mitral valve, and the interventricular septum as key anatomical features driving heart age prediction. We identified genetic (e.g titin gene) and non-genetic correlates (e.g smoking) of accelerated heart aging.

## Introduction

Cardiovascular disease [CVD] is the leading cause of death ^1^. Being an age-related disease, the prevalence of CVD is expected to starkly increase with the aging of the world population ^2,3^, leading to shortened lifespan and healthy lifespan, as well as increased healthcare cost. A strategy to reduce this burden is to further our understanding of heart aging, which can be achieved using the concept of heart age. In contrast to chronological age, which is the time that has passed since an individual was born, heart age is a measure of the changes that have accumulated in the individual’s heart over their lifetime. We refer to individuals whose heart age is higher than their chronological age as accelerated heart agers. Identifying the shared genetic and environmental factors between accelerated heart agers and between decelerated heart agers can suggest therapies and lifestyle modifications to slow or reverse heart aging.

Age-related heart changes can be leveraged to build a heart age predictor by training machine learning algorithms, such as neural networks, on heart health data to predict the participants chronological age. Age-related cardiovascular changes include stiffening of the aorta and the other vessels, fibrosis, thickening of the left ventricular free and septal walls, decrease in the end systolic and diastolic volumes, prolonged systolic contraction and diastolic relaxation and thickening and stiffening of the valves ^4–7^. After the model has successfully learned to predict chronological age using these features, the predicted age for a new participant can be interpreted as their heart age.

Attia et al. have for example built a heart age predictor on electrocardiograms [ECGs] ^8^. Similarly, ECGs and heart MRIs have also been used to predict survival ^9,10^, and a different definition of “heart age” was proposed using the Framingham cohort ^11^. Age predictors were also built for other organs, such as the brain ^12^.

In the following, we trained deep learning models on heart MRI videos, ECGs and heart health biomarkers from 45,000 UK Biobank participants aged 45-81 years to share a new light on the complex, multidimensional etiology of heart aging. In particular, we are to our knowledge the first to leverage heart MRI videos to predict heart age. We compared anatomical (MRI-based) and electrical (ECG-based) heart aging, and used attention maps to identify the anatomical and physiological features driving the prediction. We estimated the heritability of heart aging, along with SNPs associated with accelerated heart aging. Finally, we report biomarkers, clinical phenotypes, diseases, environmental and socioeconomic biomarkers associated with accelerated heart aging.

## Results

### We predicted chronological age within three years

We trained neural networks on heart MRI videos, heart MRI images (Figure 1B), electrocardiograms (Figure 1C) and heart scalar biomarkers from 45,000 UK Biobank participants (Figure 1A) aged 45-81 years (Figure S1) to build chronological age predictors (Figure 1D). We then hierarchically ensembled the models built on the different heart dimensions and subdimensions (Figure 1A) to predict chronological age with a testing root mean squared error [RMSE] 2.83±0.02 years and a R-Squared [R^2^] of 85.6±0.2% (Figure 2). We found that heart MRI videos explained more than twice the variance in chronological age compared to ECGs (R^2^= 85.3±0.1% vs. 37.6±0.4%, Figure 2), and that heart MRI videos outperformed heart MRI images as chronological age predictors (e.g. for 4-chamber view, R^2^= 78±0.2% vs. 70.5-70.9%, Figure 2). Preprocessing the images with the contrast filter had negligible effect on prediction performance. We found accelerated heart anatomical (MRI-based) and electrical (ECG-based) aging to be largely uncorrelated (Pearson correlation=.249±.005). Additionally, we predicted sex from heart MRI videos with a testing AUC-ROC of 99.8%.

**Figure 1:**
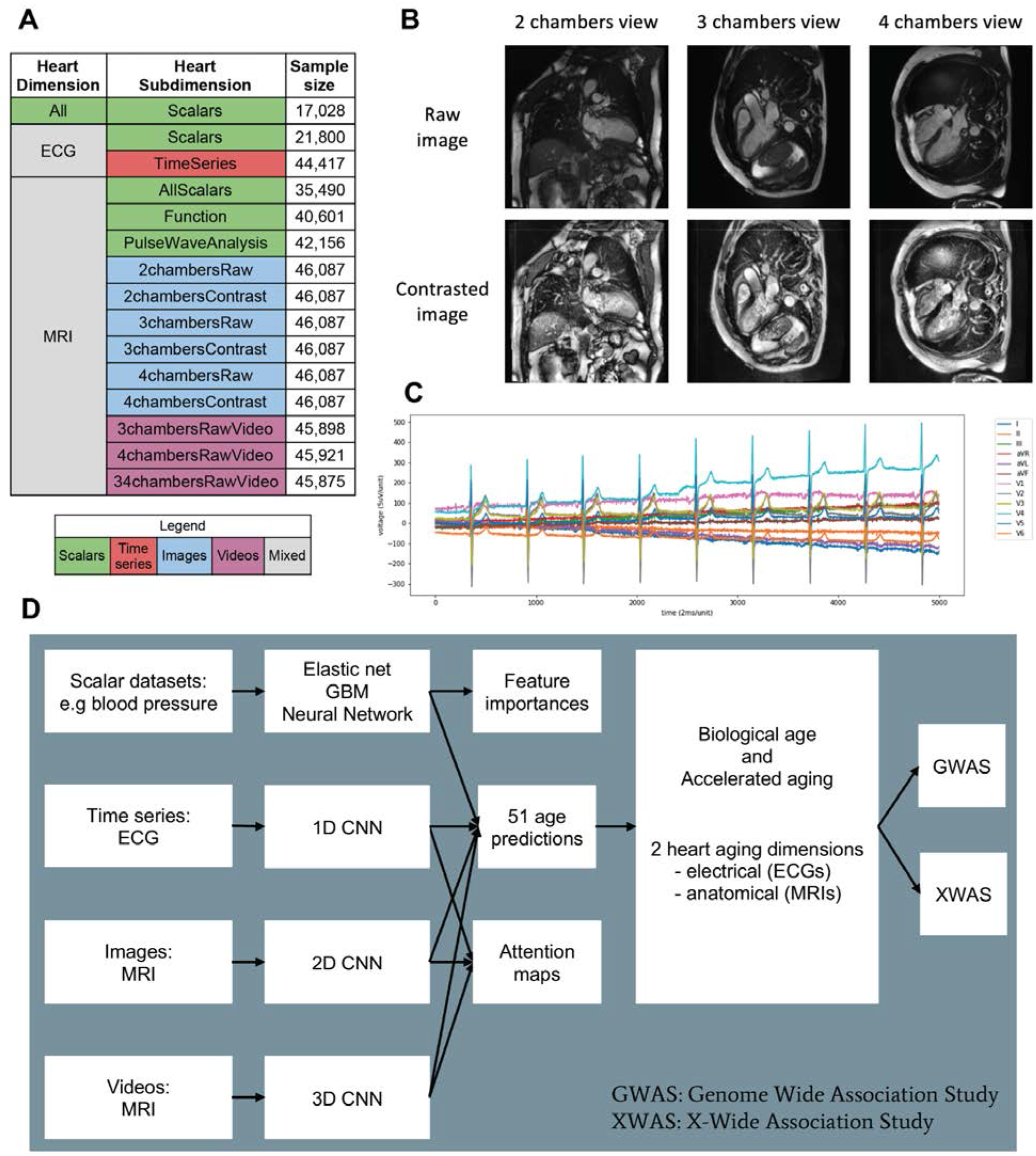
Presentation of the datasets and overview of the analytic pipeline A - UK Biobank datasets leveraged to predict chronological age organized into heart “dimensions”. B - Sample heart MRI images. C - Sample electrocardiogram. D - Overview of analytic pipeline Leftmost includes the subdimension, second left the algorithmic procedure, 3-4 columns include outputs or processed findings (such as attention maps).

**Figure 2:**
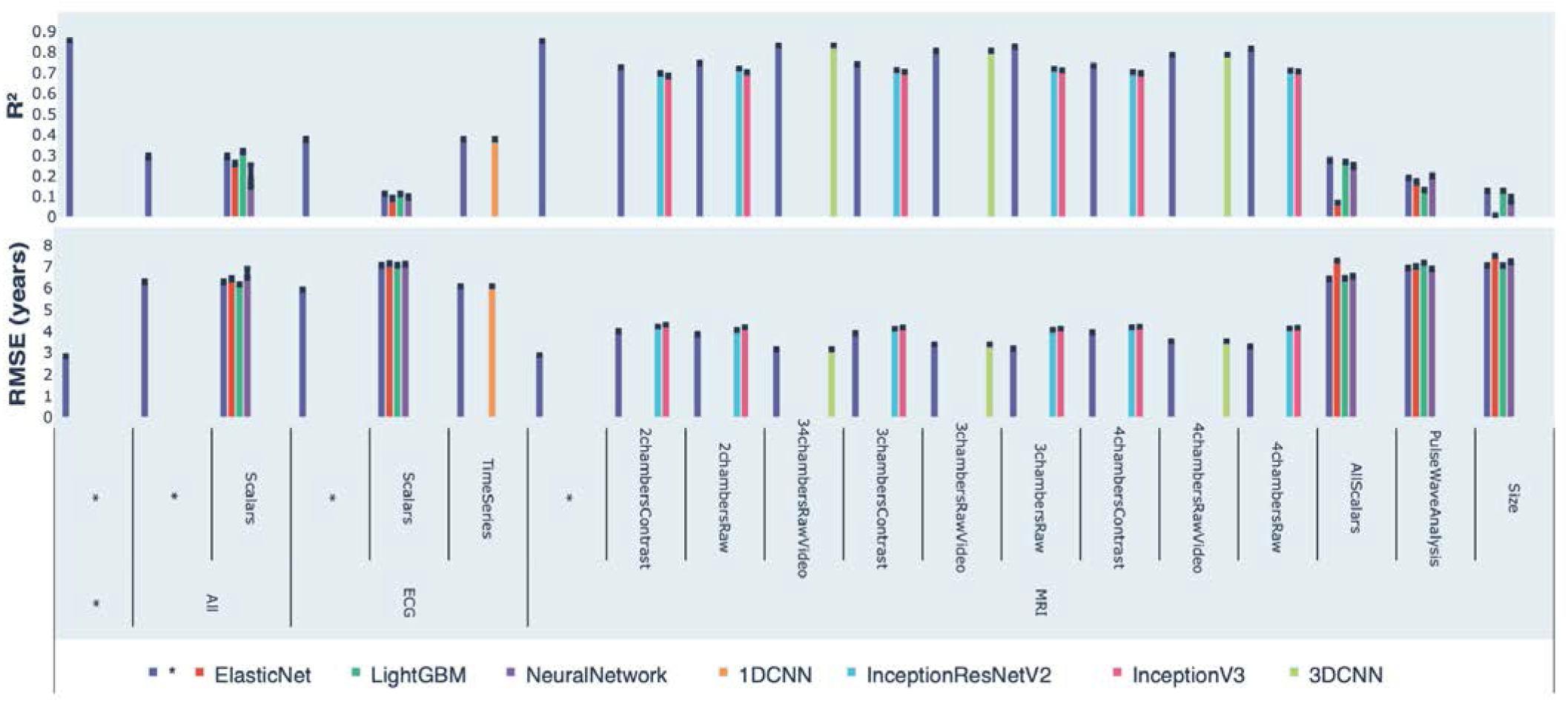
Chronological age prediction performance (R^2^ and RMSE)

### Identification of features driving heart age prediction

We used attention maps to identify the anatomical features present in the MRI videos driving the predictions of heart age. They highlighted the mitral and tricuspid valves, the aorta and the interventricular septum (Figure 3). The features driving the prediction were time-dependent and changed throughout the cardiac cycle (Figure S2).

**Figure 3:**
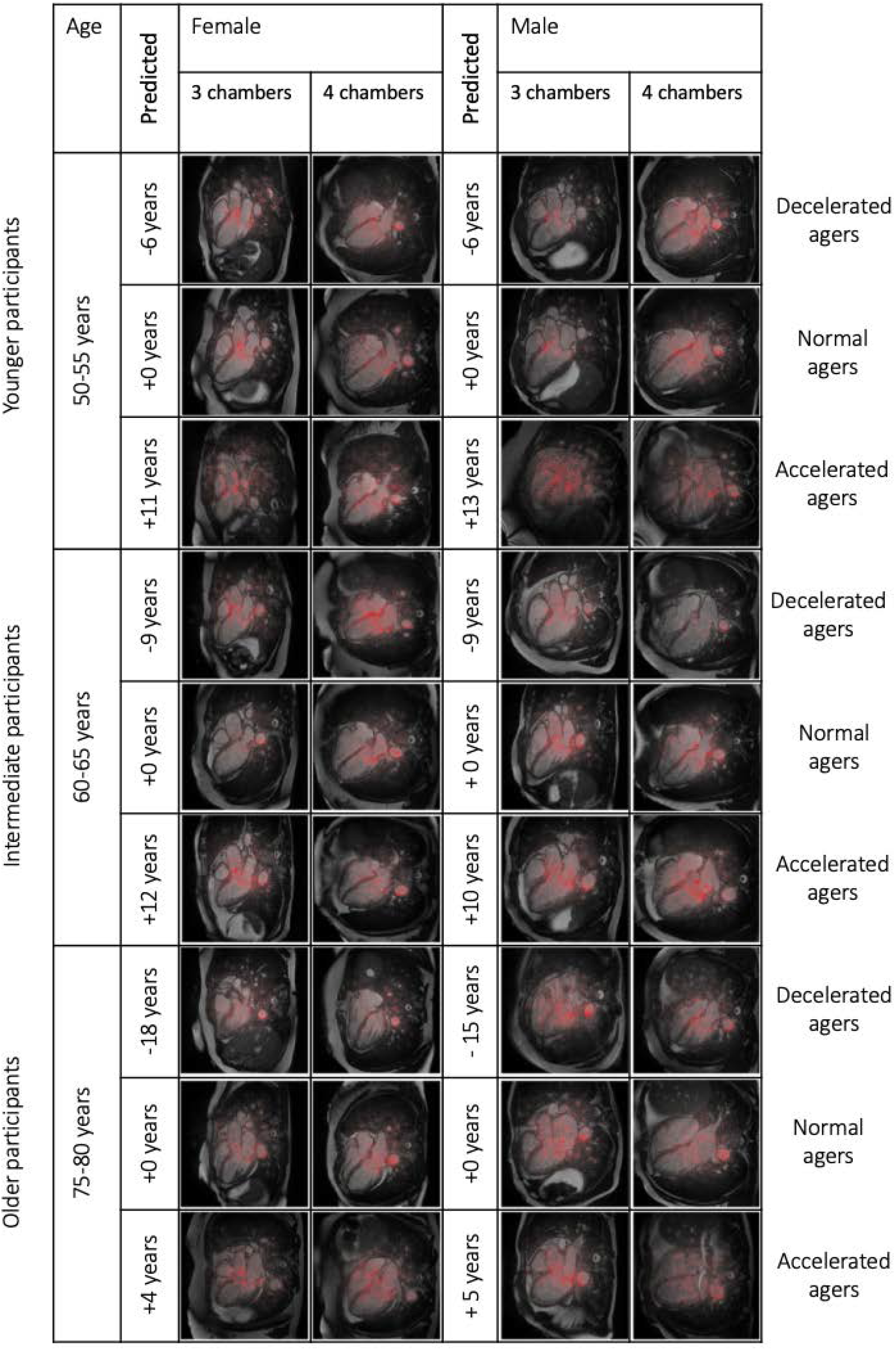
Sample attention maps for heart MRI videos.

Similarly, we used attention maps to highlight important features in MRI images (Figure S3 and Figure S4) and electrocardiograms (Figure S5). For scalar biomarkers, we found that the three most important predictors of accelerated heart anatomical aging are mean arterial pressure, systolic blood pressure and diastolic blood pressure. Similarly, the three most important predictors of accelerated heart electrical aging are the R axis, the T axis and the length of the QTC interval.

### Genetic factors and heritability of accelerated heart aging

We performed three genome wide association studies [GWASs] to estimate the GWAS-based heritability of general (h_g^2^=35.2±3.6%), anatomical (h_g^2^=37.9±1.2%), and electrical heart aging (h_g^2^=21.8±2.9%). We found accelerated anatomical and electrical heart aging to be genetically .508±.089 correlated.

We identified 24 single nucleotide polymorphisms [SNPs] in four genes associated with heart anatomical aging (Figure 4). The most significant locus is in TTN (Titin), a gene coding for a protein present in striated muscle and linked to dilated cardiomyopathy and myofibrillar myopathy). The three other genes are RP11-3N13.2 (a long intergenic non-coding RNA), RP11-224P11.1 (a pseudogene) and MASP1 (Mannan Binding Lectin Serine Peptidase 1, involved in lectin pathway of complement activation). Similarly, we identified SNPs associated with general and electrical heart aging (Figure S6 and Figure S7).

**Figure 4:**
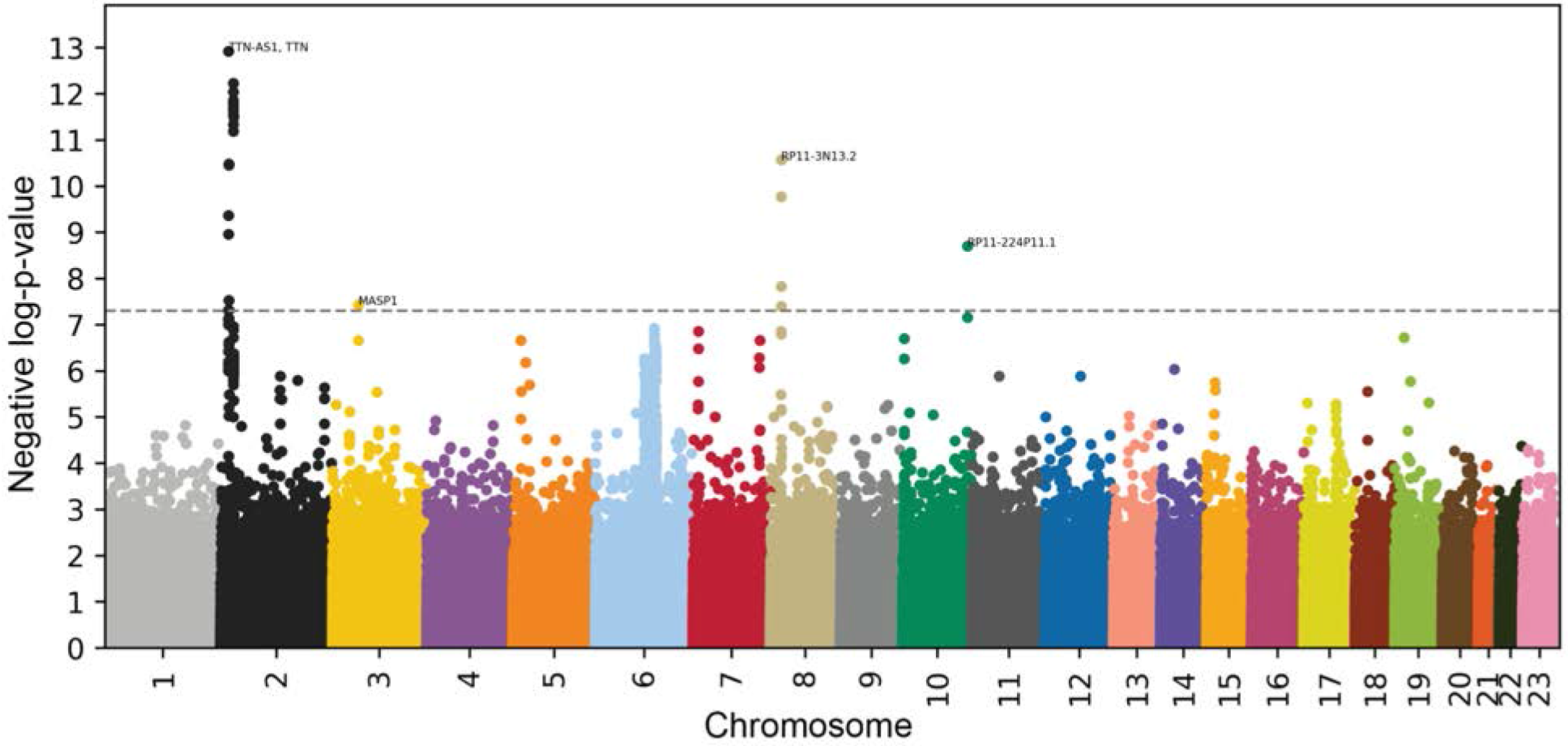
Genome Wide Association Study results negative log10(p-value) vs. chromosomal position. Dotted line denotes 5e-8.

### Non-genetic factors associated with accelerated heart aging

We performed X-wide association studies [XWASs] to identify biomarkers (Table S1), clinical phenotypes (Table S4), diseases (Table S7), environmental (Table S11) and socioeconomic (Table S14) variables associated with accelerated heart general, anatomical and electrical aging. We summarize our findings for accelerated heart anatomical aging below, because of the larger sample size leading to more significant associations. Please refer to the supplementary tables (Table S2, Table S3, Table S5, Table S6, Table S8, Table S9, Table S12, Table S13, Table S15, Table S16) for a summary of non-genetic factors associated with general, anatomical and electrical aging. The full results can be exhaustively explored at https://www.multidimensionality-of-aging.net/xwas/univariate_associations.

### Biomarkers associated with accelerated heart aging

The three biomarker categories most associated with accelerated heart aging are body impedance, blood pressure and carotid impedance. Specifically, 100.0% of anthropometry biomarkers are associated with accelerated heart aging, with the three largest associations being right leg impedance (correlation=.047), left leg impedance (correlation=.044), and whole body impedance (correlation=.044). 100.0% of blood pressure biomarkers are associated with accelerated heart aging, with the three largest associations being higher levels of diastolic blood pressure (correlation=.166), systolic blood pressure (correlation=.158), and pulse rate (correlation=.082). 100.0% of carotid ultrasound biomarkers are associated with accelerated heart aging, with the three largest associations being mean carotid intima-medial thickness at 150, 240 and 120 degrees (respective correlations of .054, .051 and .050).

Conversely, the three biomarkers categories most associated with decelerated heart aging are higher levels of spirometry (an indicator of lung function), higher ability in cognitive digit substitution and higher hand grip strength. Specifically, the three spirometry biomarkers (forced expiratory volume in one second, peak exploratory flow, forced vital capacity) are associated with decelerated heart aging (respective correlations of .083, .063, .040). The three symbol digits substitutions (a cognitive test) biomarkers are associated with decelerated heart aging (number of symbol digit matches made correctly: correlation=.042. Number of symbol digit matches attempted: correlation=.042). The two hand grip strength biomarkers are associated with decelerated heart aging (left and right hand grip correlations are respectively .054 and .048).

### Clinical phenotypes associated with accelerated heart aging

The three clinical phenotype categories most associated with accelerated heart aging are chest pain, breathing and claudication. Specifically, 100.0% of chest pain clinical phenotypes are associated with accelerated heart aging, with the three largest associations being chest pain and discomfort (correlation=.033), chest pain or discomfort walking normally (correlation=.031), and chest pain ceases when standing still (correlation=.031). The two breathing clinical phenotypes are associated with accelerated heart aging (shortness of breath walking on level ground: correlation=.057, wheeze or whistling in the chest in the last year: correlation=.035). 46.2% of claudication variables are associated with accelerated heart aging, with the three largest associations being leg pain when walking uphill or hurrying (correlation=.032), leg pain on walking (correlation=.030) and leg pain when walking normally (correlation=.029).

Conversely, the three clinical phenotypes categories most associated with decelerated heart aging are sexual factors (age first had sexual intercourse: correlation=.040), cancer screening (most recent bowel cancer screening: correlation=.044) and mouth health (no mouth/teeth dental problems: correlation=.029).

### Diseases associated with accelerated heart aging

The three disease categories most associated with accelerated heart aging are cardiovascular diseases, general health, and mental disorders. Specifically, 10.4% of cardiovascular diseases are associated with accelerated heart anatomical aging, with the three largest associations being hypertension (correlation=.095), atrial fibrillation and flutter (.065) and chronic ischaemic heart disease (correlation=.040). 8.3% of general health variables are associated with accelerated heart anatomical aging, with the three largest associations being personal history of disease (correlation=.056), personal history of medical treatment (correlation=.056) and problems related to lifestyle (correlation=.042).

The only disease categories associated with decelerated heart anatomical aging are related to obstetrics (e.g. outcome of delivery, correlation=.037).

### Environmental variables associated with accelerated heart aging

The three environmental variables categories most associated with accelerated heart anatomical aging are smoking, alcohol and sun exposure. Specifically, 37.5% of smoking variables are associated with accelerated heart anatomical aging with the three largest associations being pack years adult smoking as proportion of life span exposed to smoking (correlation=.112), pack years of smoking (correlation= .109), and past tobacco smoking: smoked on most or all days (correlation=.083). 13.8% of alcohol variables are associated with accelerated heart anatomical aging, with the three largest associations being average weekly beer plus cider intake (correlation=.042), average weekly spirits intake (.031) and average weekly champagne plus white wine intake (correlation=.029). 10.0% of sun exposure variables are associated with accelerated heart anatomical aging with the two associated variables being facial aging: about your age (correlation=.032) and facial aging: older than you are (correlation=.029).

Conversely, the three environmental variable categories most associated with decelerated heart anatomical aging are physical activity, smoking and sleep. Specifically, 40.0% of physical activity variables are associated with decelerated heart anatomical aging, with the strongest associations being frequency of strenuous sports in the last four weeks (correlation=.077), duration of strenuous sports (.074) and frequency of other exercises in the last four weeks (correlation=.068). 29.2% of smoking variables are associated with decelerated heart anatomical aging with the three largest associations being smoking status: never (correlation=.077), age started smoking in former smokers (correlation=.076) and current tobacco smoking: no (correlation=.059). In terms of sleep variables, snoring is associated with decelerated heart aging (correlation=.041).

### Socioeconomic variables associated with accelerated heart aging

The three socioeconomic variables categories most associated with accelerated heart anatomical aging are social support, sociodemographics and employment. Specifically, 22.2% of social support variables are associated with accelerated heart anatomical aging, with the two associated variables being leisure/social activities: pub or social club (correlation=.038) and leisure/social activities: none (correlation=.025). In terms of sociodemographics, accelerated heart anatomical aging is associated with private healthcare (correlation=.024), and in terms of employment, it is associated with current employment status: retired (correlation=.065).

Conversely, the three socioeconomic variables categories most associated with decelerated heart anatomical aging are employment, sociodemographics and education. Specifically, 30.4% of employment variables are associated with accelerated heart anatomical aging, with the three largest associations being length of working week for main job (correlation=.056), current employment status: in paid employment or self-employed (correlation=.054), and time employed in main current job (correlation=.035). In terms of sociodemographics variables, accelerated heart anatomical aging is associated with not receiving attendance/disability/mobility allowance (correlation=.035), and in terms of education, it is associated with qualifications: college or university degree (correlation=.035).

## Discussion

### Novelty and strength

Our composite heart age predictor is the first to leverage heart MRI videos, as well as the first to combine information from both MRI, electrocardiograms and scalar biomarkers. We predicted chronological age with high accuracy (R^2^=85.6±0.2%, RMSE=2.81±0.02 years), significantly outperforming previous heart age predictors (R^2^ ∼ 70%)^8^. We also claim that there is a biological basis of heart aging across the diverse dimensions queried here, with strong heritability and associations with environmental factors.

### Comparison between heart anatomical and electrical agings

Heart anatomical (MRI-based) and electrical (ECG-based) agings are two distinct facets of heart aging. Although largely phenotypically uncorrelated (correlation=.249±.002), we found that they share important genetic factors, as shown by their genetic correlation (.508±.089). The hypothesis that MRI videos and ECGs capture distinct but related facets of heart aging is further supported by the association between biomarkers and accelerated heart anatomical versus electrical aging. Both anatomical and electrical aging are associated with blood pressure and ECG biomarkers, but ECG biomarkers are more strongly associated with electrical aging, whereas blood pressure biomarkers are more strongly associated with anatomical aging.

### Utility of heart age predictors

Biological age predictors can be used to quantify the effect of environmental effects or rejuvenating drugs candidates ^13^. For example, the DNA methylation clock is being used in a clinical trial to evaluate the efficiency of NAD+ as a rejuvenating therapy ^14^, and was used to show that obesity is associated with accelerated liver aging ^15^. Aging is multidimensional, and the predictions outputted by different biological age predictors tend to be weakly correlated, measuring different facets of aging ^16,17^. Heart age predictors such as ours are therefore particularly well suited to evaluate the effects of rejuvenating therapies on cardiovascular health, and provide complementary information to other methods such as the Framingham risk score ^11^.

### Genetic and non-genetic factors associated with heart aging

We found that accelerated heart aging is GWAS-heritable (35.2±3.6%) and associated with SNPs in four genes (e.g Titin, linked to heart diseases), which represent potential therapeutic targets to slow heart aging.

Accelerated heart aging is associated with cardiovascular biomarkers such as blood pressure, carotid ultrasound, and arterial stiffness measurements, as well as non-cardiovascular biomarkers such as brain MRI features, cognitive tests, muscular strength, bone health, spirometry, blood and urine biochemistry. We observed a similar pattern in terms of association with clinical phenotypes and diseases. Accelerated heart aging is associated not only with cardiovascular phenotypes (e.g chest pain) and diseases (e.g hypertension, atrial fibrillation and flutter, ischaemic heart disease, tachycardia), but also with non-cardiovascular phenotypes (e.g mental health, claudication, eyesight, mouth health) and diseases (respiratory diseases, metabolic disorders, mental disorders, cancer), suggesting that heart aging is correlated with aging of other organ systems. More generally, the association between heart aging and general health (e.g general history of disease/medical treatment, overall health rating, long-standing illness, disability or infirmity) suggests that, although distinct from aging in other organs, heart aging is influenced by a “general aging” factor. Interestingly, accelerated heart aging is for example associated with perceived age (looking younger/older than one’s age). We further explore the connection between heart aging and aging in different organ systems in a different paper ^16^.

We identified lifestyle factors associated with accelerated heart aging (e.g smoking, drinking, physical activity, diet), partially aligned with the existing literature ^18^ and suggesting potential lifestyle interventions to slow or reverse heart aging. We stress that correlation is not causation and that the aforementioned are only candidate lifestyle changes.

Another example of association between accelerated heart aging and environmental exposures is taking a cholesterol lowering medication, which is associated with accelerated heart aging. Causality here, in contrast with time-invariant genetics, should not be misinterpreted. Taking the medication likely does not accelerate heart aging. Instead, accelerated heart aging might be caused by poor cardiovascular health, including high cholesterol levels, hence the prescription of a cholesterol lowering medication. A future direction would be to test whether patients whose heart MRI video was recorded twice, once before and once after starting cholesterol lowering medication (or a suite of hypothesized rejuvenating therapies), to examine heart rejuvenation compared to participants who did not. More generally, leveraging the subset of UKB participants with longitudinal data to identify lifestyle interventions and medications associated with a reduction in heart age would allow us to get closer to establishing causality.

Socioeconomic status is a complex variable, measured and assessed in different ways, including education and income. However measured, it is an important health determinant. In the US, the richest 1% males live on average 14.6±0.2 years longer than the poorest 1% males, and the richest 1% females live on average 10.1±0.2 years longer than the poorest 1% females^19^. To the degree that the increased lifespan of individuals who are fortunate who have higher socioeconomic status can be captured by our new characterization of heart aging, we found that higher socioeconomic status (e.g holding a university degree, income, number of vehicles) is associated with decelerated heart aging, likely due to better healthcare and health literacy ^20^.

We found that being comparatively taller at age ten is associated with accelerated heart aging in adulthood. However, height is reported to be negatively correlated with blood pressure ^21^. We explain our findings by the fact that height and blood pressure are positively correlated (∼.4) at age twenty, with the correlation progressively decreasing with age and reaching ∼-.2 at age eighty. Specifically, in the UK Biobank cohort, the correlation becomes negative around sixty-five years ^22^. With the majority of UKB participants being younger than 70 years old (Figure S1), tall participants have been disproportionately exposed to the effects of high blood pressure, explaining their accelerated heart aging.

### Anatomical heart features driving age prediction

The attention maps showed that the three-dimensional convolutional neural network relies on the mitral and tricuspid valves, the aorta and the interventricular septum to predict heart anatomical age. The thickness of the interventricular septum increases with age ^23^, whose sigmoid-shape is part of normal aging ^24^. The attention maps also highlighted the mitral and tricuspid valves, which undergo significant changes with age ^25^, such as calcification. Calcification is rare for the tricuspid valve ^26^, but common for the mitral one ^27^. The attention maps highlighted the aorta as well, whose composition, stiffness, thickness and diameter change with age ^28,29^. More generally, the high age prediction accuracy obtained (R2=85.3+-0.1%) can be explained by the numerous changes affecting the cardiovascular system with age, as reported in the introduction. For example, atherosclerosis develops, stiffening the arterial vessels and forcing the heart to pump harder ^30^. As another example, heart rate decreases with age in the UKB cohort (correlation=-.048±.018), contrary to what was previously reported in the literature ^31^. Finally, the image-based attention maps showed that the models did not exclusively rely exclusively on heart features to predict chronological age, but also on the surrounding tissue and anatomical features.

### Leveraging raw electrocardiogram time series improved prediction accuracy

In terms of ECG-based models, we found that leveraging the raw time series significantly improved the prediction performance compared to relying on the sole scalar biomarkers (R^2^=37.6±0.4% vs. 11.1±0.4%). Our model outperformed Attia et al.’s model ^8^ in terms of absolute error (RMSE= 6.09±0.02 years vs. MAE=6.9±5.6 years) but underperformed in terms of R^2^ (37.6±0.4% vs. 70%), which can likely be explained by a difference in age range (45-81 vs. 18-100 years) and sample size (21,792 vs. 275,056).

### The cardiac cycle contains information that is not captured by a single MRI image

The models trained on MRI videos significantly outperformed the models trained on MRI images (R^2^= 78±0.2% vs. 70.5-70.9%), despite the 3D-CNN architecture being significantly simpler and shallower than the 2D-CNN architectures we transfer learned from (3D CNN: five layers vs. InceptionResNetv2: 164 layers). Additionally, the 3D-CNN models trained on MRI videos had only access to 150*150 pixels time-frame images, as opposed to 200*200 pixels images for the 2D-CNN models, and could therefore not rely as much on non-cardiovascular features present in the surrounding tissue. An example of anatomical features that could be better leveraged by models built on MRI videos is the closing of the valves. While images-based models also highlight this feature, they cannot measure at which point in the cycle the valves are closing, or how long they remain closed. We conclude from the higher performance of video-based models that the temporal component of the cardiac cycle encodes important information regarding heart aging, and we recommend that future projects leverage full MRI videos as opposed to sample images, even if this decision must be taken at the expense of the complexity of the convolutional architecture used.

A potential advantage of image-based over video-based models is that image-based models only require that the full cardiac cycle was recorded to calculate the participant’s biological age, which is costlier. However, participant MRI images collected need to match the step in the heart cycle on which the image-based models were trained, or prediction accuracy will be lower (Figure S8).

### Age predictors suffer from a bias

We observed a bias: participants on the younger end of the age range tend to be predicted older than they are, and the participants on the older end of the age range tend to be predicted younger than they are (Figure 2), observed tentatively in other cohorts ^8^. This bias is not biological in nature. For example, splitting the age range into two groups (the “young” and the “old”) and retraining a model on each half yields a similar pattern. In this experiment, on each half, the youngest participant will be predicted older, and the older participants will be predicted younger, independently of their absolute chronological age. This suggests that the problem is statistically driven. We found that the bias increases when the performance of the model decreases, and that a similar pattern can be observed when predicting other numerical phenotypes, such as blood pressure. Our interpretation is that age predictions are biased towards the mean age of the age range distribution as this is how the model minimizes R^2^ in the absence of information. As more information is collected by the model, it’s performance improves and this bias is reduced, but it still behaves as if a prior centered around the mean age had been specified.

### Predicting age vs. predicting survival

Age predictors can be built by training machine learning models to predict age-related phenotypes. such as chronological age and survival. Predicting chronological age and survival are two ways to estimate biological age, but they have their differences. Some individuals could maintain a youthful body for the duration of their life, but die suddenly if one of the vital organs quickly fails. In contrast, some individuals could show early signs of aging but survive for a long time as long as their vital organs function at a sufficient level. The advantage of defining biological age as a function of survivability is that it is a hard, easily measurable and concrete outcome, unlike age. The drawback is that survival does not reflect the progression of aging. Between 15 years old and 44 years old, the probability of dying from a non-traumatic event is very low, so a survival model will not be able to efficiently capture the aging process. In contrast, chronological age is not a phenotype in itself, but it is a strong proxy for both biological age, age related diseases and survival, and it is easier to follow and track even in the absence of the onset of a specific disease or a death event, for example in early and middle adulthood. Leveraging our pipeline to predict survival and the onset of age-related cardiovascular diseases could allow us to discover other facets of heart aging. A survival predictor has already been generated on a different dataset ^10^.

### Limitations

While the attention maps and the associations between our predictor and both genetic and non-genetic factors suggest that the age prediction is driven by cardiovascular features (aorta, mitral valve, interventricular septums), some of the attention maps also highlight other anatomical features in the surrounding tissue. To improve the biological relevance of the model and better capture heart aging rather than general thoracic aging, a possibility would be to first use image segmentation (manually or automatically) to extract the heart segment, before retraining the models on this subset of the images.

Different datasets and cohorts can use different MRI machines and settings, which can limit the transferability of the model trained on the sole UK Biobank dataset. Leveraging other datasets would allow us to both estimate the out-dataset generalization error and train more generalizable models.

To reduce the size of each sample, we used every other time frame in the time dimension, which decreased the total amount of information available. While 25 time frames might still be enough to efficiently describe the cardiac cycle, it is plausible that training a model on the full 50 time frames videos would lead to a gain of performance.

Finally, the UKB is a cross-sectional, observational dataset, and the correlations we observed between accelerated heart aging and non-genetic factors do not allow us to infer causation.

## Methods

### Data and materials availability

We used the UK Biobank (project ID: 52887). The code can be found at https://github.com/Deep-Learning-and-Aging. The results can be interactively and extensively explored at https://www.multidimensionality-of-aging.net/. We will make the biological age phenotypes available through UK Biobank upon publication. The GWAS results can be found at https://www.dropbox.com/s/59e9ojl3wu8qie9/Multidimensionality_of_aging-GWAS_results.zip?dl=0.

### Software

Our code can be found at https://github.com/Deep-Learning-and-Aging. For the genetics analysis, we used the BOLT-LMM ^32,33^ and BOLT-REML ^34^ softwares. We coded the parallel submission of the jobs in Bash ^35^.

### Cohort Dataset: Participants of the UK Biobank

We leveraged the UK Biobank^36^ cohort (project ID: 52887). The UKB cohort consists of data originating from a large biobank collected from 502,211 de-identified participants in the United Kingdom that were aged between 37 years and 74 years at enrollment (starting in 2006). Out of these participants, 45,000 had their electrocardiogram and a heart MRI video collected (Figure 1A). The Harvard internal review board (IRB) deemed the research as non-human subjects research (IRB: IRB16-2145).

### Definition of the different heart dimensions

We defined heart aging dimensions by hierarchically grouping the different datasets at two different levels: we call the first level “heart dimensions” and the second level “heart subdimensions”. The complete hierarchy between the three heart dimensions and the 15 subdimensions is described in Figure 1A. For example, one of the heart dimensions we investigated was electrical heart age, whose estimation is ECG-based. This dimension consists of two subdimensions: (1) the scalar features (the length of the different intervals, such as PP, PQ and QT), and (2) the time series (the raw 12 leads ECG signal recorded for 600 timesteps). Similarly, “MRI”, which refers to heart anatomical aging, encompasses 12 subdimensions that include three based on scalar data (e.g., Size, Pulse Wave Analysis), six of them based on MRI images (e.g., 2chambersContrast), and three of them based on MRI videos (e.g. 3chambersRawVideo). Finally, the “All” dimension encompasses all the scalar biomarkers from both the ECG and the MRI dimensions.

### Data types and Preprocessing

The data preprocessing step is different for the different data modalities: demographic variables, scalar predictors, time series, images and videos. We define scalar predictors as predictors whose information can be encoded in a single number, such as blood pressure, as opposed to data with a higher number of dimensions such as time series (one dimension, which is time), images (two dimensions, which are the height and the width of the image) and videos (three dimensions, which are the height and width of the video, along with time).

### Demographic variables

First, we removed out the UKB samples for which age or sex was missing. For sex, we used the genetic sex when available, and the self-reported sex when genetic sex was not available. We computed age as the difference between the date when the patient attended the assessment center and the year and month of birth of the patient to estimate the patient’s age with greater precision. For physical activity, the age of the patient when they attended the assessment center was different from the age of the patient when they wore the wrist accelerometer, so we computed their age based on the date they started wearing the device. For ECG, we found 26 samples for which the date at which the participant attended the assessment center was not recorded in the main data frame, so we imputed it using data from the ECG files. We one-hot encoded ethnicity.

### Scalar data

We define scalar data as a variable that is encoded as a single number, such as height or blood albumin level, as opposed to data with a higher number of dimensions, such as time series, images and videos. The complete list of scalar biomarkers can be found in Table S1 under “Heart”. We did not preprocess the scalar data, aside from the normalization that is described under cross-validation further below.

### Time series: electrocardiograms

The UKB contains 44,463 10 second-long 12-lead resting electrocardiograms [ECGs] (field 20205), stored as .xml files. We extracted the raw signal from the .xml files using the Beautiful soup python package ^37^ (Figure S9) along with the participants’ heart rate. We then used the heart rate to compute the length of every participant’s heart cycle. We used this information to decompose each ECG into four components called the “level” (the mean value), the “trend” (the linear bias over time), the “seasonality” (the periodical component of the signal) and the “noise” (the allegedly random remaining variation), using the seasonal_decompose function from the statsmodel python library ^38^ (Figure S10). To generate our signal, we subtracted the trend component from the raw signal. We filtered out the samples for which the heart rate was lower than 30 beats per minute or higher than 125 beats per minute, as we found by observing the ECGs that these outlying values were associated with corrupted signal (Figure S11). After completing the preprocessing, we were left with 44,417 samples.

During our preliminary analysis, we preprocessed the data in two other ways. These preprocessing methods were outperformed by the preprocessing described above, so we did not use them for our final pipeline, but we briefly describe them here as other researchers might find them of interest. First, we use the median ECG signal for each participant. Unlike the ten-second-long signal described above, this signal is a single heartbeat for each participant, stored as 600 time steps. It can be directly extracted from the .xml files provided by the UKB. Second, we split each ten-second-long ECG signal into several single beat signals. To do this, we extracted the heart rate from the .xml for each sample, we used it to compute the length of a cardiac cycle for the participant, and we split the sample accordingly. This generated approximately ten times more samples for each participant.

### Videos: MRI

The UKB contains long-axis heart MRI videos (field 20,208, 46,869 samples for 44,413 participants), stored as 50 time 200*200 DICOM images. The 50 images do not correspond to a specific sampling frequency. Pictures are instead collected over several cardiac cycles (where a cycle is one heartbeat). They are selected and rearranged until the full cardiac cycle of the participant can be reconstituted. As a consequence, two participants with two different heart rates will still have their full cardiac cycle summarized by 50 images. We filtered out samples for which the number of time frames was different from 50.

Three different views of the heart were recorded by UKB: (1) two-chamber view (displaying the right atrium and ventricle), (2) three-chamber views (displaying the two atria and the left ventricle) and (3) four-chamber view (displaying the two atria and ventricles). We filtered out 844 samples for which one or more of the three views was not available. We applied a histogram equalizer filter to enhance the contrast of the images (similarly to what we did for the liver and pancreas images). We stored both the raw and the contrasted images, which we respectively refer to as “Raw” and “Contrast”.

For the images, we saved the first frame of each of the remaining samples or the remaining samples without cropping the images, as mentioned under “Preprocessing - Images - Heart”. For the videos, we cropped the images to obtain 150*150 images centered on the heart, with the aim to reduce the size of the model needed to train on the videos. For this same reason, we generated two 25 time frames-long samples from each 50 time frames video: one with the even numbered time frames, and one with the odd numbered time frames. This last step allowed us to halve the size of each image, therefore doubling the batch size that would fit into the model. Because our preliminary results on images showed a lesser predictive potential for the two-chamber view, we only trained 3D CNN models on three-chamber view and four chamber view videos.

### Heart MRI Images

We selected the first time frame of the videos before the cropping step and converted them into RGB pictures by duplicating the grayscale layer. We obtained 46,987 images of dimensions 200*200*3 out of the initial grayscale videos of dimensions 200*200*50. We rescaled the values of the pixels to be between 0 and 1 instead of between 0 and 255 to mitigate the explosion of gradients in the neural networks (see Methods-Algorithms). Starting from grayscale images, we saved them as RGB by duplicating the values three times. As explained in detail under Methods-Algorithms-Images, we used transfer learning to analyze the images. In the context of transfer learning, the size of the architecture of the model is affected by the dimension of the input. Large images contain more information but require larger deep learning architectures that take longer to train. To resolve prohibitory long training times, we resized the images so that the total number of pixels for each channel would be below 100,000 (316*316 pixels). A sample of preprocessed images can be found in Figure S12. All the preprocessed images were stored as .jpg.

### Data augmentation

#### Images

To prevent overfitting and increase our sample size during the training we used data augmentation ^39^ on the images. Each image was randomly shifted vertically and horizontally (10%), as well as rotated (10 degrees). We chose the hyperparameters for these transformations’ distributions to represent the variations we observed between the images in the initial dataset.

The data augmentation process is dynamically performed during the training. Augmented images are not generated in advance. Instead, each image is randomly augmented before being fed to the neural network for each epoch during the training.

#### Videos

For the heart MRI videos, we introduced a random rotation of plus or minus three degrees in the spatial plane, sampled from the uniform distribution [-3,3]. Similarly to images, the augmentation process for videos is performed dynamically during the training. Each video is randomly augmented before being fed to the neural network.

### Machine learning algorithms

For scalar datasets, we used elastic nets, gradient boosted machines [GBMs] and fully connected neural networks. For times series, images and videos we used one-dimensional, two-dimensional, and three-dimensional convolutional neural networks, respectively. We describe these models in detail in the Supplementary.

### Scalar data

We used three different algorithms to predict age from scalar data (non-dimensional variables, such as laboratory values). Elastic Nets [EN] (a regularized linear regression that represents a compromise between ridge regularization and LASSO regularization), Gradient Boosted Machines [GBM] (LightGBM implementation ^40^), and Neural Networks [NN]. The choice of these three algorithms represents a compromise between interpretability and performance. Linear regressions and their regularized forms (LASSO ^41^, ridge ^42^, elastic net ^43^) are highly interpretable using the regression coefficients but are poorly suited to leverage non-linear relationships or interactions between the features and therefore tend to underperform compared to the other algorithms. In contrast, neural networks ^44,45^ are complex models, which are designed to capture non-linear relationships and interactions between the variables. However, tools to interpret them are limited ^46^ so they are closer to a “black box”. Tree-based methods such as random forests ^47^, gradient boosted machines ^48^ or XGBoost ^49^ represent a compromise between linear regressions and neural networks in terms of interpretability. They tend to perform similarly to neural networks when limited data is available, and the feature importances can still be used to identify which predictors played an important role in generating the predictions. However, unlike linear regression, feature importances are always non-negative values, so one cannot interpret whether a predictor is associated with older or younger age. We also performed preliminary analyses with other tree-based algorithms, such as random forests ^47^, vanilla gradient boosted machines ^48^ and XGBoost ^49^. We found that they performed similarly to LightGBM, so we only used this last algorithm as a representative for tree-based algorithms in our final calculations.

### Time series: Electrocardiograms

We used the architecture from the article “Prediction of mortality from 12-lead electrocardiogram voltage data using a deep neural network” ^9^. The original architecture can be found under “Extended Data Fig. 2” in the original paper.

The architecture of our model can be found in Figure S13 and Figure S14. Our architecture is composed of two blocks: (1) the convolutional block and (2) the dense block. The convolutional block comprises five seven-layer deep one-dimensional convolutional neural networks branches, in parallel. Each of these CNN branches takes three ECG leads of 5,040 time steps (ten seconds, we padded 20 time steps before and after the raw signal to mirror the original model ^9^) as its input, with some of the leads being used as input for more than one CNN. We used zero-padding for the convolutional layers. Each CNN’s branch’s output is processed by a max pooling to convert the 512 one-dimensional filters into 512 scalar features, which are then concatenated with 64 features built on sex and ethnicity. These 2,624 features are then inputted into the dense block, which is a six-layer-deep fully connected neural network. The number of nodes in the first four layers is halved every layer (256, 128, 64, 32). The fifth layer has eight nodes, and the sixth layer has a single node with a linear activation function to predict age. We applied batch normalization, kernel and bias regularization, and we used ReLU as activation function for every layer in the architecture, except for the final layer.

In other words, we made the following changes to adapt the original architecture to the task of predicting chronological age instead of one year survival. (1) We replaced chronological age by ethnicity as a side predictor. (2) We replaced the final sigmoid activation layer by a linear activation layer. (3) We used the same architecture for the five CNN branches, unlike the original architecture which is using different CNN architectures for the different CNN branches. (4) We used zero-padding on the convolutional layers which conserved their input’s dimension. We relied on max pooling layers with a size of two and a stride of two to iteratively halve the input’s size. (5) We added batch normalization and ReLU as activation functions to the dense layers, except for the final dense layer (see (2)).

During our preliminary analysis, we explored simpler architectures using one-dimensional convolutional layers, LSTM layers or GRU layers. We found that our architectures did not perform as well (custom one-dimensional CNNs: R^2^=30%; LSTMs and GRUs: R^2^=20%) as the architecture described above so we did not train these models for our final pipeline. In particular, we found that LSTM and GRU models significantly underperformed, with R^2^ values lower than 10%.

We used Adam ^50^ as the compiler for all the time series models.

### Images

#### Convolutional Neural Networks Architectures

We used transfer learning ^51–53^ to leverage two different convolutional neural networks ^54^ [CNN] architectures pre-trained on the ImageNet dataset ^55–57^ and made available through the python Keras library ^58^: InceptionV3 ^59^ and InceptionResNetV2 ^60^. We considered other architectures such as VGG16 ^61^, VGG19 ^61^ and EfficientNetB7 ^62^, but found that they performed poorly and inconsistently on our datasets during our preliminary analysis and we therefore did not train them in the final pipeline. For each architecture, we removed the top layers initially used to predict the 1,000 different ImageNet images categories. We refer to this truncated model as the “base CNN architecture”.

We added to the base CNN architecture what we refer to as a “side neural network”. A side neural network is a single fully connected layer of 16 nodes, taking the sex and the ethnicity variables of the participant as input. The output of this small side neural network was concatenated to the output of the base CNN architecture described above. This architecture allowed the model to consider the features extracted by the base CNN architecture in the context of the sex and ethnicity variables. For example, the presence of the same anatomical heart feature can be interpreted by the algorithm differently for a male and for a female. We added several sequential fully connected dense layers after the concatenation of the outputs of the CNN architecture and the side neural architecture. The number and size of these layers were set as hyperparameters. We used ReLU ^63^ as the activation function for the dense layers we added, and we regularized them with a combination of weight decay ^64,65^ and dropout ^66^, both of which were also set as hyperparameters. Finally, we added a dense layer with a single node and linear activation to predict age.

#### Compiler

The compiler uses gradient descent ^67,68^ to train the model. We treated the gradient descent optimizer, the initial learning rate and the batch size as hyperparameters. We used mean squared error [MSE] as the loss function, root mean squared error [RMSE] as the metric and we clipped the norm of the gradient so that it could not be higher than 1.0 ^69^.

We defined an epoch to be 32,768 images. If the training loss did not decrease for seven consecutive epochs, the learning rate was divided by two. This is theoretically redundant with the features of optimizers such as Adam, but we found that enforcing this manual decrease of the learning rate was sometimes beneficial. During training, after each image has been seen once by the model, the order of the images is shuffled. At the end of each epoch, if the validation performance improved, the model’s weights were saved.

We defined convergence as the absence of improvement on the validation loss for 15 consecutive epochs. This strategy is called early stopping ^70^ and is a form of regularization. We requested the GPUs on the supercomputer for ten hours. If a model did not converge within this time and improved its performance at least once during the ten hours period, another GPU was later requested to reiterate the training, starting from the model’s last best weights.

### Videos

Our architecture is largely inspired by the C3D ^71^ architecture. We treated time as a third spatial dimension and built a three-dimensional convolutional neural network [3D CNN] to analyze it. The architecture of the model can be found in Table S40 and is composed of a convolutional block and a dense block. The convolutional block consists of four three-dimensional convolutional layers. The first layer has 16 cubic kernels of dimensions 3×3×3, the second has 64 cubic kernels of dimensions 3×3×3, the third has 512 cubic kernels of dimensions 5×5×5, and the fourth layer has 1,024 kernels of dimensions 5×5×7, the third dimension being time. All convolutional layers have a stride of 1×1×1 and are zero padded to conserve the dimension of their input. We progressively reduce the spatial dimensions of the input sample by using a max pooling operation with dimension 2×2×1 and stride 2×2×1 after each convolutional layer. The fifth layer is a max-pooling layer in the spatial dimensions. The features are then flattened and batch normalized ^72^ before being fed to the dense block. The dense block consists of two dense layers. The penultimate layer is a fully connected layer with 1,024 nodes with dropout. The final layer is a fully connected layer with a single node with a linear activation to predict chronological age. All other layers use SELU ^73^ as their activation function.

We used the same compiler for the videos as for the images (see Methods - Machine learning algorithms - Images - Compiler), with the exception that an epoch is defined as seeing each sample once, and that the learning rate is decreased by 20% if the training performance does not improve for five consecutive epochs.

We also built a CNN-LSTM architecture on the heart MRI videos. The CNN extracts spatial features from each frame of the video. These spatial features are then fed as a time series as input to the LSTM that leverages the temporal dimension. We selected the 2D CNN architecture that performed best on heart MRI images and we used it to extract 64 features from each image. This step gave 64 50 time frames-long time series for each sample video. We then built an LSTM on this dataset to predict chronological age. We found that the CNN-LSTM architecture underperformed compared to the 3D CNN architecture (R^2^=69.8±0.9%), so we did not include it in our final pipeline.

### Training, tuning and predictions

We split the entire dataset into ten data folds. We then tuned the models built on scalar data, on time series, on images and on videos using four different pipelines. For scalar data-based models, we performed a nested-cross validation. For time series-based, images-based and video-based models, we manually tuned some of the hyperparameters before performing a simple cross-validation. We describe the splitting of the data into different folds and the tuning procedures in greater detail in the Supplementary.

### Interpretability of the machine learning predictions

To interpret the models, we used the regression coefficients for the elastic nets, the feature importances for the GBMs, a permutation test for the fully connected neural networks, and attention maps (saliency and Grad-RAM) for the convolutional neural networks (Supplementary Methods).

### Ensembling to improve prediction and define aging dimensions

We built a three-level hierarchy of ensemble models to improve prediction accuracies. At the lowest level, we combined the predictions from different algorithms on the same aging sub-subdimension. For example, we combined the predictions generated by the elastic net, the gradient boosted machine and the neural network on the ECG (dimension) scalar predictors (subdimension). At the second level, we combined the predictions from different subdimensions of a unique dimension. For example, we combined the ECG subdimensions “Scalars” and “Time Series” into an ensemble prediction. Finally, at the highest level, we combined the predictions from the three heart aging dimensions into a general heart age prediction. The ensemble models from the lower levels are hierarchically used as components of the ensemble models of the higher models. For example, the ensemble models built by combining the algorithms at the lowest level for each of the MRI sub-subdimensions are leveraged when building the general heart aging ensemble model.

We built each ensemble model separately on each of the ten data folds. For example, to build the ensemble model on the testing predictions of the data fold #1, we trained and tuned an elastic net on the validation predictions from the data fold #0 using a 10-folds inner cross-validation, as the validation predictions on fold #0 and the testing predictions on fold #1 are generated by the same model (see Methods - Training, tuning and predictions - Images - Scalar data - Nested cross-validation; Methods - Training, tuning and predictions - Images - Cross-validation). We used the same hyperparameters space and Bayesian hyperparameters optimization method as we did for the inner cross-validation we performed during the tuning of the non-ensemble models.

To summarize, the testing ensemble predictions are computed by concatenating the testing predictions generated by ten different elastic nets, each of which was trained and tuned using a 10-folds inner cross-validation on one validation data fold (10% of the full dataset) and tested on one testing fold. This is different from the inner-cross validation performed when training the non-ensemble models, which was performed on the “training+validation” data folds, so on 9 data folds (90% of the dataset).

### Evaluating the performance of models

We evaluated the performance of the models using two different metrics: R-Squared [R^2^] and root mean squared error [RMSE]. We computed a confidence interval on the performance metrics in two different ways. First, we computed the standard deviation between the different data folds. The test predictions on each of the ten data folds are generated by ten different models, so this measure of standard deviation captures both model variability and the variability in prediction accuracy between samples. Second, we computed the standard deviation by bootstrapping the computation of the performance metrics 1,000 times. This second measure of variation does not capture model variability but evaluates the variance in the prediction accuracy between samples.

### Biological age definition

We defined the biological age of participants as the prediction generated by the model corresponding to heart dimension or subdimension, after correcting for the bias in the residuals.

We indeed observed a bias in the residuals. For each model, participants on the older end of the chronological age distribution tend to be predicted younger than they are. Symmetrically, participants on the younger end of the chronological age distribution tend to be predicted older than they are. This bias does not seem to be biologically driven. Rather it seems to be statistically driven, as the same 60-year-old individual will tend to be predicted younger in a cohort with an age range of 60-80 years, and to be predicted older in a cohort with an age range of 60-80. We ran a linear regression on the residuals as a function of age for each model and used it to correct each prediction for this statistical bias.

After defining biological age as the corrected prediction, we defined accelerated aging as the corrected residuals. For example, a 60-year-old whose MRI data predicted an age of 70 years old after correction for the bias in the residuals is estimated to have a biological heart anatomical age of 70 years, and an accelerated heart anatomical aging of ten years.

It is important to understand that this step of correction of the predictions and the residuals takes place after the evaluation of the performance of the models but precedes the analysis of the biological ages properties.

### Genome-wide association of accelerated heart aging

The UKB contains genome-wide genetic data for 488,251 of the 502,492 participants^74^ under the hg19/GRCh37 build.

We used the average accelerated aging value over the different samples collected over time (see Supplementary - Models ensembling - Generating average predictions for each participant). Next, we performed genome wide association studies [GWASs] to identify single-nucleotide polymorphisms [SNPs] associated with heart accelerated general, anatomical and electrical agings using BOLT-LMM ^32,33^ and estimated the the SNP-based heritability for each of our biological age phenotypes, and we computed the genetic pairwise correlations between dimensions using BOLT-REML ^34^. We used the v3 imputed genetic data to increase the power of the GWAS, and we corrected all of them for the following covariates: age, sex, ethnicity, the assessment center that the participant attended when their DNA was collected, and the 20 genetic principal components precomputed by the UKB. We used the linkage disequilibrium [LD] scores from the 1,000 Human Genomes Project ^75^. To avoid population stratification, we performed our GWAS on individuals with White ethnicity.

### Identification of SNPs associated with accelerated aging

We identified the SNPs associated with accelerated heart aging dimensions and subdimensions using the BOLT-LMM ^32,33^ software (p-value of 5e-8). The sample size for the genotyping of the X chromosome is one thousand samples smaller than for the autosomal chromosomes. We therefore performed two GWASs for each aging dimension. (1) excluding the X chromosome, to leverage the full autosomal sample size when identifying the SNPs on the autosome. (2) including the X chromosome, to identify the SNPs on this sex chromosome. We then concatenated the results from the two GWASs to cover the entire genome, at the exception of the Y chromosome.

We plotted the results using a Manhattan plot and a volcano plot. We used the bioinfokit ^76^ python package to generate the Manhattan plots. We generated quantile-quantile plots [Q-Q plots] to estimate the p-value inflation as well.

### Heritability and genetic correlation

We estimated the heritability of the accelerated aging dimensions using the BOLT-REML ^34^ software. We included the X chromosome in the analysis and corrected for the same covariates as we did for the GWAS. Using the same software and parameters, we computed the genetic correlations between accelerated heart anatomical and electrical agings.

We annotated the significant SNPs with their matching genes using the following four steps pipeline. (1) We annotated the SNPs based on the rs number using SNPnexus ^77–81^. When the SNP was between two genes, we annotated it with the nearest gene. (2) We used SNPnexus to annotate the SNPs that did not match during the first step, this time using their genomic coordinates. After these two first steps, 30 out of the 9,697 significant SNPs did not find a match. (3) We annotated these SNPs using LocusZoom ^82^. Unlike SNPnexus, LocusZoom does not provide the gene types, so we filled this information with GeneCards ^83^. After this third step, four genes were not matched. (4) We used RCSB Protein Data Bank ^84^ to annotate three of the four missing genes. One gene on the X chromosome did not find a match (position 56,640,134).

### Non-genetic correlates of accelerated aging

We identified non-genetically measured (i.e factors not measured on a GWAS array) correlates of each aging dimension, which we classified in six categories: biomarkers, clinical phenotypes, diseases, family history, environmental, and socioeconomic variables. We refer to the union of these association analyses as an X-Wide Association Study [XWAS]. (1) We define as biomarkers the scalar variables measured on the participant, which we initially leveraged to predict age (e.g. blood pressure, Table S1). (2) We define clinical phenotypes as other biological factors not directly measured on the participant, but instead collected by the questionnaire, and which we did not use to predict chronological age. For example, one of the clinical phenotypes categories is eyesight, which contains variables such as “wears glasses or contact lenses”, which is different from the direct refractive error measurements performed on the patients, which are considered “biomarkers” (Table S4). (3) Diseases include the different medical diagnoses categories listed by UKB (Table S7). (4) Family history variables include illnesses of family members (Table S10). (5) Environmental variables include alcohol, diet, electronic devices, medication, sun exposure, early life factors, medication, sun exposure, sleep, smoking, and physical activity variables collected from the questionnaire (Table S11). (6) Socioeconomic variables include education, employment, household, social support and other sociodemographics (Table S14). We provide information about the preprocessing of the XWAS in the Supplementary Methods.

## Data Availability

https://www.multidimensionality-of-aging.net/

https://www.dropbox.com/s/59e9ojl3wu8qie9/Multidimensionality_of_aging-GWAS_results.zip?dl=0

## Acknowledgments

We would like to thank Dr. Yassine Moustarhfir for helping us interpret the heart MRI images and videos, Raffaele Potami from Harvard Medical School research computing group for helping us utilize O2’s computing resources, and Pauline Dimaano for proofreading the manuscript. We also want to acknowledge UK Biobank for providing us with access to the data they collected. The UK Biobank project number is 52887. We thank HMS RC for computing support.

Funding: NIEHS R00 ES023504; NIEHS R21 ES25052; NIAID R01 AI127250; NSF 163870; MassCATS, Massachusetts Life Science Center; Sanofi. The funders had no role in the study design or drafting of the manuscript(s).

## Author Contributions

**Alan Le Goallec:** (1) Designed the project. (2) Supervised the project. (3) Predicted chronological age from heart MRI images. (4) Computed the attention maps for heart MRI images. (4) Generated preliminary results for the chronological age predictors built on ECGs. (5) Ensembled the models, evaluated their performance, computed biological ages and estimated the correlation structure between the aging dimensions. (6) Performed the genome wide association studies. (5) Designed the website. (6) Wrote the manuscript.

**Jean-Baptiste Prost:** (1) Preprocessed the datasets for heart MRI videos. (2) Predicted chronological age from heart MRI videos. (3) Computed the attention maps for videos.

**Sasha Collin:** (1) Preprocessed the ECG time series. (2) Predicted chronological age using the ECG time series datasets. (3) Computed the attention maps for the time series.

**Samuel Diai:** (1) Predicted chronological age from scalar features. (2) Wrote the python class to build an ensemble model using a cross-validated elastic net. (3) Performed the X-wide association study.

**Théo Vincent:** (1) Website data engineer. (2) Improved and polished the website, with a particular emphasis on the XWAS section.

**Chirag J. Patel:** (1) Supervised the project. (2) Edited the manuscript. (3) Provided funding.

